# PREDICT breast v4.0: An update to the PREDICT breast prognostic model

**DOI:** 10.1101/2025.08.28.25334663

**Authors:** Paul D.P. Pharoah, Yi-Wen Hsiao, Gordon C. Wishart, Pei-Chen Peng

## Abstract

The PREDICT breast prognostic and treatment benefit model has undergone several revisions since its first release. The most recent version (v3.1) was developed using a data set of 35,474 cases diagnosed between 2000 and 2017 in a single region of England. PREDICT breast provides predicted outcomes at 5, 10 and 15 years, but most clinicians use the 10-year outcomes for decision making. The purpose of this study was to reparameterize the model using a larger data set from across the UK and to compare the performance of v4.0 with that of v3.1.

There were 172,208 eligible cases randomly split 50:50 into model development and validation data sets. Cox proportional hazards models were derived for estrogen receptor negative and estrogen receptor positive cancer for breast cancer specific mortality with a third model for non-breast cancer mortality.

In cases with at least five years follow-up and censored at ten years, the model was well-calibrated with a less than 5% difference between observed and predicted breast cancer deaths. Model discrimination was also good with AUCs in the validation data of 0.735 and 0.794 for ER negative and ER positive cases respectively. Calibration and discrimination were slightly improved compared to PREDICT breast v3.1.

## Introduction

The PREDICT breast cancer prognostication and treatment benefit prediction model (v1) was developed in 2010 using data from the UK East Anglia Cancer Registration and Information Centre (ECRIC) for model fitting and data from the West Midlands Cancer Intelligence Unit for model validation ^1-3^. PREDICT v1 was implemented as a web-based tool for clinicians in January 2011 (www.breast.predict.nhs.uk), and since then the use of the tool has increased steadily around the world. The model was refitted in 2017 using the original cohort of cases from East Anglia with updated survival time in order to take into account age at diagnosis and to smooth out the hazard ratio functions for tumour size and node status (v2) ^4^ and the website was redesigned (now at https://breast.predict.cam). A further update using a larger, more contemporary data set was published in 2023 (v3). PREDICT breast v3 was developed using data from 4,644 ER-negative and 30,830 ER-positive breast cancer cases diagnosed from 2000 to 2017 in the region served by the Eastern Cancer Registry in the UK (available at https://breast.v3.predict.cam). PREDICT has been independently validated in cohorts from Canada ^5^, India^6^, Japan^7^, Malaysia ^8^, the Netherlands ^9-11^, New Zealand^12^, Spain^13^, the UK ^14 15^ and the USA^16-18^ and has generally been shown to have good discrimination and calibration. PREDICT breast provides predicted outcomes at 5, 10 and 15 years after diagnosis, but for clinical decision making most clinicians use the 10-year outcomes. The purpose of this analysis was to use a larger data set to re-parameterize the model to optimize performance in the most recently diagnosed cases.

## Methods

### Patient data

All analyses were carried out on anonymized data from the National Cancer Registration and Analysis Service for all women diagnosed in England with non-metastatic invasive breast cancer from 2000 to 2017 inclusive. The use of these data complies with all relevant ethical and data governance regulations. Patient information is collected by NCRAS without the need for patient consent under Section 251 of the National Health Service Act 2006 and its current Regulations, the Health Service (Control of Patient Information) Regulations 2002. Ethical approval by the National Research Ethics Service was not required because all analyses were carried out on an anonymized data set.

Information on all cases included age at diagnosis, year of diagnosis, tumour size, histological grade, tumour stage at diagnosis, number of lymph nodes sampled, number of lymph nodes positive, ER status, HER2 status, mode of detection (clinically detected vs. screen detected), and whether the patient had undergone chemotherapy, hormone therapy and/or radiotherapy within 6 months of diagnosis. Patients younger than 25 or older than 85 at diagnosis, patients with a tumour larger than 20 centimetres, or with more than 20 positive lymph nodes were excluded from the analysis. Of 372,110 cases, complete data for age at diagnosis, year of diagnosis, tumour size, histological grade, tumour stage at diagnosis, number of lymph nodes sampled, number of lymph nodes positive, ER status were available for 172,708 (46%). These were randomly split 50:50 into a model training data set and a model testing data set. The sample sizeof the training data was more than sufficient (compared with the ∼35,000 cases used in the original v3 development cohort) to derive stable parameter estimates, while preserving a similarly large independent validation set for unbiased performance assessment.

Details of the specific regimen used for radiotherapy, chemotherapy, duration of hormonal therapy, or use of trastuzumab or bisphosphonates were not available. We assumed that all patients that underwent chemotherapy were treated with an anthracycline-based regimen and that all women received hormonal therapy for five years. The benefits of radiotherapy were applied to all patients who received it including those who had lumpectomy and those who had mastectomy as the primary surgical treatment. Death certificate flagging through the Office for National Statistics provides the registries with notification of deaths. The lag times for these are a few weeks for cancer deaths and 2 months to 1 year for non-cancer deaths. Vital status was ascertained at the end of December 2019, and so all analyses were censored on 31 December 2018 to allow for delay in reporting of vital status. Breast cancer-specific mortality was defined as deaths where breast cancer was listed as the cause of death on part 1a, 1b or 1c of the death certificate.

### Statistical methods

The general approach to model development was similar to that use for the development of v2 and v3. Multivariable Cox proportional hazards models were used to estimate the prognostic effect of each variable. In all models follow up time was defined as the time from breast cancer diagnosis to last follow up, death or 15 years after diagnosis, whichever came first. The outcome of interest was either breast cancer-specific mortality or mortality from other causes. Separate models were derived for breast cancer-specific mortality in ER-negative and ER-positive cases. Multivariable fractional polynomials were used to model non-linear effects between the continuous risk factors (age at diagnosis, tumour size and number of positive nodes) and breast cancer-specific mortality to improve the fit to the data in the presence of non-linearity. Sequential backward elimination with a maximum of 4 degrees of freedom for a single continuous predictor was used to estimate the continuous variable transformations. Age at diagnosis was transformed to age at diagnosis minus 24 so that the baseline hazard corresponds to that of a case diagnosed age 24. Year of diagnosis was grouped into before 2011 and 2011 and later; the latter was used as the baseline category so that the baseline hazard function will be that for more contemporary patients. Parameter estimates for HER2 ^2^, KI67 ^3^ and PR ^12^ for previous versions of PREDICT breast were derived from external data. There was a substantial proportion of missing data for these variables and so the same parameter estimates were used for version 4.0.

The relative treatment benefits for chemotherapy, hormone therapy and radiotherapy were constrained to the estimates of benefit from the randomised controlled trial meta-analyses of the Early Breast Cancer Trialists Collaborative Group (adjuvant hormone therapy log hazard ratio -0.386 ^19^, adjuvant chemotherapy log hazard ratio -0.248 ^20^, radiotherapy log hazard ratio -0.180 ^21^) by adding them as an offset in the breast cancer specific mortality analyses. The relative harms of chemotherapy and radiotherapy were constrained to the estimates of benefit reported by Kerr and colleagues (adjuvant chemotherapy log hazard ratio 0.183)^22^ and Taylor and colleagues (radiotherapy log hazard ratio 0.078 per Gray whole-heart dose)^23^ by adding them as an offset in the analysis of other mortality.

After fitting the Cox proportional hazards models to ER-negative and ER-positive cases, a multiple fractional polynomial model with a Gaussian distribution was fit to the baseline hazards according to the method of Sauberei and colleagues to derive smoothed baseline hazard functions for breast cancer-specific mortality and non-breast cancer specific mortality.

## Results

A total of 172,708 cases were included in the analyses. The characteristics of the cases in the training and validation data sets by ER status are shown in Table 1. Cox proportional hazards models were built using multi-variable fractional polynomials. The fractional polynomial functions for the variables of interest are shown in Table 2 together with the associated hazard ratios and 95% confidence limits. The hazard ratio as a function of the fractional polynomials for age at diagnosis, tumour size and number of positive nodes are shown in Figure 1. The observed baseline hazards and the fitted curve for the corresponding multi-variable fractional polynomial regression models are shown in Figure 2.

**Table 1:** Characteristics of female breast cancer cases by ER status.

|  | ER negative |  | ER positive |  |
| --- | --- | --- | --- | --- |
|  | Training data | Test data | Training data | Test data |
| Number | 11,767 | 11,820 | 74,231 | 74,890 |
| Age diagnosis, n (%) |  |  |  |  |
| <40 | 848 ( 7.2) | 843 ( 7.1) | 2,606 ( 3.5) | 2,539 ( 3.4) |
| 40-59 | 4,930 (41.9) | 4,917 (41.6) | 31,619 (42.6) | 32,177 (43.0) |
| 60-69 | 3,051 (25.9) | 3,053 (25.8) | 23,294 (31.4) | 23,448 (31.3) |
| >=70 | 2,938 (25.0) | 3,007 (25.4) | 16,712 (22.5) | 16,726 (22.3) |
| Grade, n (%) |  |  |  |  |
| 1 | 199 ( 1.7) | 176 ( 1.5) | 14,822 (20.0) | 14,719 (19.7) |
| 2 | 2,279 (19.4) | 2,264 (19.2) | 42,152 (56.8) | 42,493 (56.7) |
| 3 | 9,289 (78.9) | 9,380 (79.4) | 17,257 (23.2) | 17,678 (23.6) |
| Tumour size, n (%) |  |  |  |  |
| 1-19 mm | 5,156 (43.8) | 5,059 (42.8) | 42,838 (57.7) | 42,819 (57.2) |
| 20-49 mm | 5,947 (50.5) | 6,036 (51.1) | 28,080 (37.8) | 28,828 (38.5) |
| 50+ mm | 664 ( 5.6) | 725 ( 6.1) | 3,313 ( 4.5) | 3,243 ( 4.3) |
| Number of positive nodes, n (%) |  |  |  |  |
| 0 | 8,217 (69.8) | 8,090 (68.4) | 53,083 (71.5) | 53,112 (70.9) |
| 1-3 | 2,421 (20.6) | 2,501 (21.2) | 16,104 (21.7) | 16,626 (22.2) |
| 4-9 | 752 ( 6.4) | 809 ( 6.8) | 3,597 ( 4.8) | 3,747 ( 5.0) |
| 10+ | 377 ( 3.2) | 420 ( 3.6) | 1,447 ( 1.9) | 1,405 ( 1.9) |
| Mode of detection, n (%) |  |  |  |  |
| Clinical detected | 8,618 (73.2) | 8,737 (73.9) | 44,075 (59.4) | 44,681 (59.7) |
| Unknown | 732 ( 6.2) | 729 ( 6.2) | 3,991 ( 5.4) | 4,032 ( 5.4) |
| Screen detected | 2,417 (20.5) | 2,354 (19.9) | 26,165 (35.2) | 26,177 (35.0) |

**Table 2:** Fractional polynomial functions for variables in the Cox proportional hazards regression models with associated hazard ratios and 95% confidence limits.

| Variable | MFP function | Hazard ratio | LCL | UCL |
| --- | --- | --- | --- | --- |
| *ER negative model breast cancer specific mortality* |  |  |  |  |
| Age at diagnosis | $((\text{age}-24)/100)^3$ | 32.6 | 15.8 | 67.5 |
| Tumour size (mm) | $\log(\text{size}/10)$ | 2.03 | 1.86 | 2.22 |
| Number of positive nodes | $\log(\text{nodes}+1)$ | 1.78 | 1.69 | 1.87 |
| Tumour grade |  | 1.28 | 1.14 | 1.43 |
| Mode of detection |  |  |  |  |
| Missing |  | 1.01 | 0.85 | 1.20 |
| Screen detected |  | 0.65 | 0.55 | 0.76 |
| Year diagnosis before 2011 |  | 1.30 | 1.18 | 1.44 |
| *ER positive model breast cancer specific mortality* |  |  |  |  |
| Age at diagnosis 1 | $((\text{age}-24)/100)^{0.5}$ | 0.0009 | 0.0001 | 0.0076 |
| Age at diagnosis 2 | $(\text{age}-24)/100$ | 2868 | 422 | 19499 |
| Number of positive nodes | $\log(\text{nodes}+1)$ | 12.5 | 8.8 | 17.8 |
| Tumour size 1 (mm) | $(\text{size}/10)^{0.5}$ | 0.64 | 0.58 | 0.70 |
| Tumour size 2 (mm) | $(\text{size}/10)$ | 1.94 | 1.87 | 2.01 |
| Tumour rade |  | 2.12 | 2.00 | 2.24 |
| Mode of detection |  |  |  |  |
| Missing |  | 1.24 | 1.10 | 1.39 |
| Screen detected |  | 0.65 | 0.59 | 0.72 |
| Year diagnosis before 2011 |  | 1.51 | 1.40 | 1.63 |
| *Non breast cancer mortality* |  |  |  |  |
| Age at diagnosis 1 | $((\text{age}-24)/100)^{-0.5}$ | 0.54 | 0.41 | 0.72 |
| Age at diagnosis 2 | $((\text{age}-24)/100)^2$ | 28475 | 14087 | 57559 |
| Year diagnosis before 2011 |  | 1.02 | 0.95 | 1.09 |

MFP = multi-variable fractional polynomial; LCL = lower 95% confidence limit; UCL = upper 95% confidence limit

**Figure 1.**
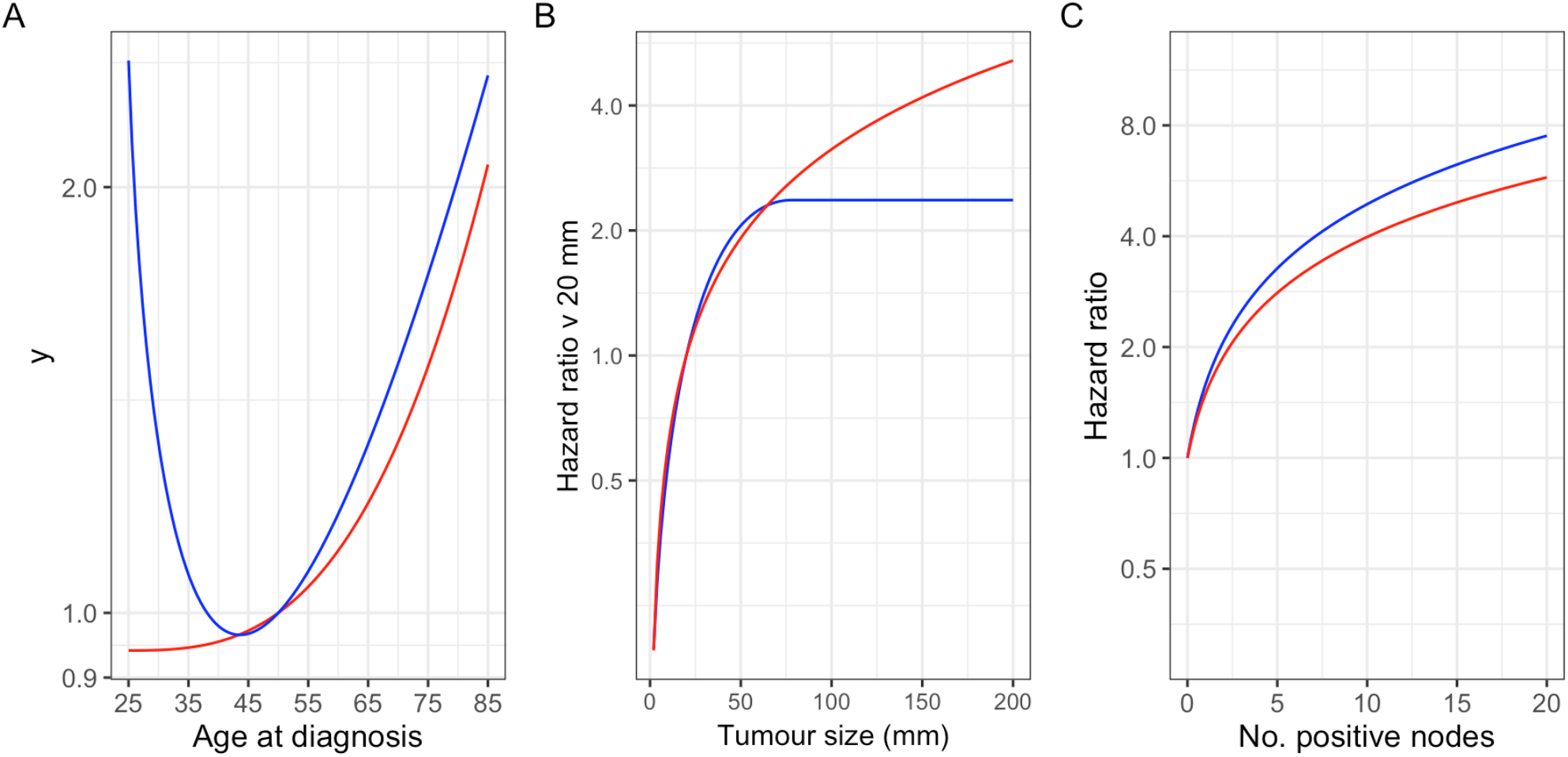
Fractional polynomial hazard ratio functions for age at diagnosis (A), tumour size (B) and number of positive lymph nodes (C). Blue line ER positive cases, red line ER negative cases.

**Figure 2.**
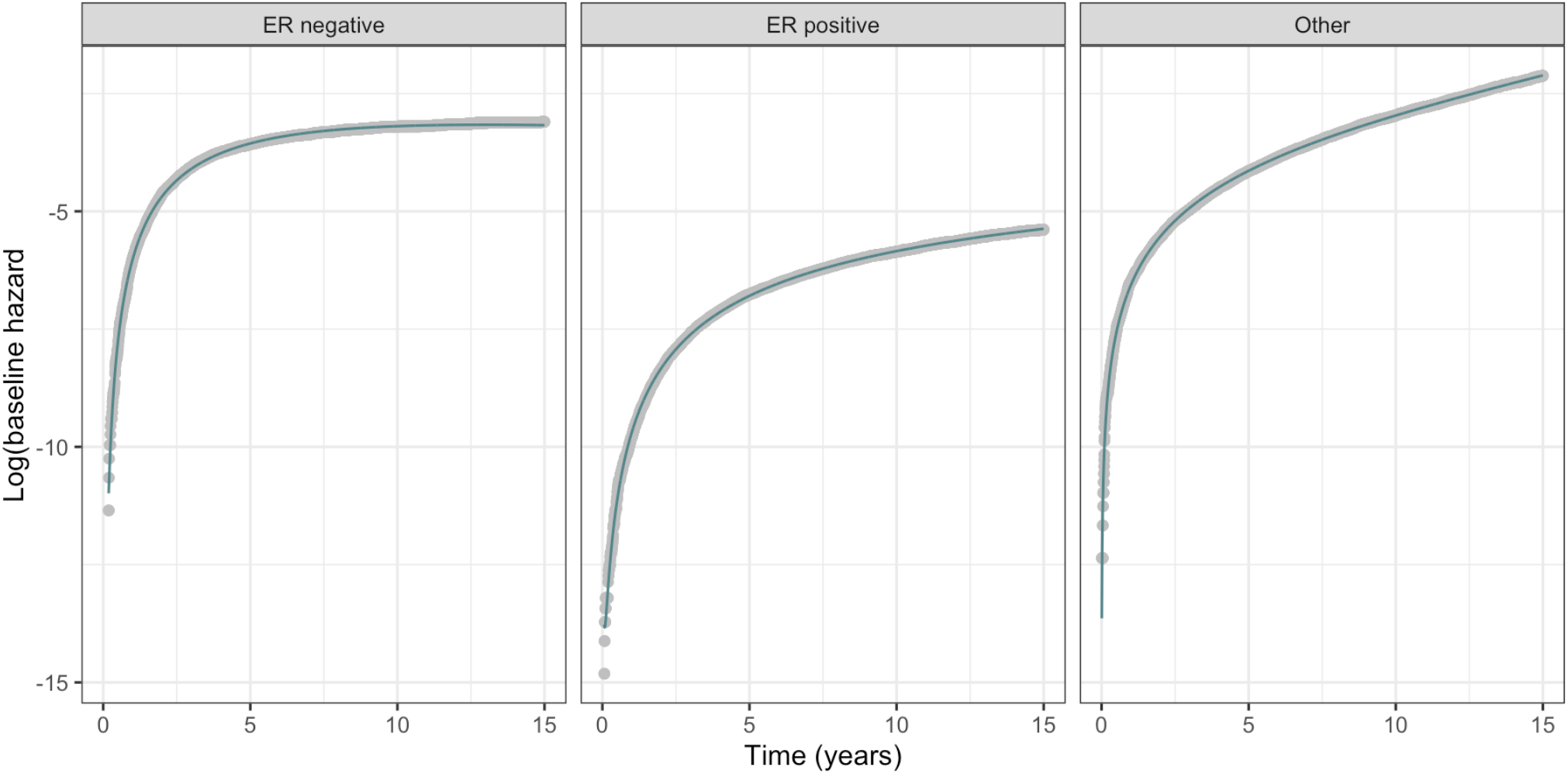
Observed baseline hazards (grey dots) and multi-variable fractional polynomial fit (blue line) for breast cancer specific mortality in ER negative cases and ER positive cases and other mortality in all cases

The model, designated PREDICT breast v4.0, based on the coefficients from the Cox proportional hazards model and the derived baseline hazard functions was applied to the subset of 56,357 cases (training data 28,010 cases; validation data 28,347 cases) diagnosed after 2010 with at least 5 years potential follow-up to obtain the expected breast mortality at 10 years. Breast cancer mortality and non-breast cancer mortality are treated as competing risks in the model. Expected breast mortality was then compared with the observed breast cancer mortality and the expected mortality based on v3.1 (Table 3). Calibration was excellent in both training and validation data with less than 5% difference between the observed and predicted deaths from breast cancer. Goodness-of-fit was evaluated by comparing observed and predicted breast deaths in tenths of predicted risk. Figure 3 shows that there was good calibration of predicted deaths across all levels of risk. Discrimination was calculated as the area under the receiver operator characteristic curve. Discrimination of the ER positive model was somewhat better than that of the ER negative model. There was a small improvement in both calibration and discrimination for v4.0 compared to v3.1.

**Table 3:** Model calibration and discrimination for PREDICT breast v4.0 compared to v3.1.

| ER status | Number of cases | Observed deaths | Predicted deaths |  |  |  | Discrimination |  |
| --- | --- | --- | --- | --- | --- | --- | --- | --- |
|  |  |  | v3.1 | Diff (%)* | v4.0 | Diff %* | v3.1 | v4.0 |
| Model development data |  |  |  |  |  |  |  |  |
| Negative | 3,802 | 739 | 725 | -1.9 | 752 | 1.8 | 0.717 | 0.717 |
| Positive | 24,208 | 1,439 | 1,534 | 6.6 | 1479 | 2.7 | 0.783 | 0.787 |
| Model validation data |  |  |  |  |  |  |  |  |
| Negative | 3,848 | 771 | 749 | -2.9 | 774 | 0.4 | 0.734 | 0.735 |
| Positive | 24,499 | 1,456 | 1,559 | 7.1 | 1,503 | 3.2 | 0.791 | 0.794 |

\* $100 \times (\text{observed} - \text{predicted}) / \text{observed}$

**Figure 3.**
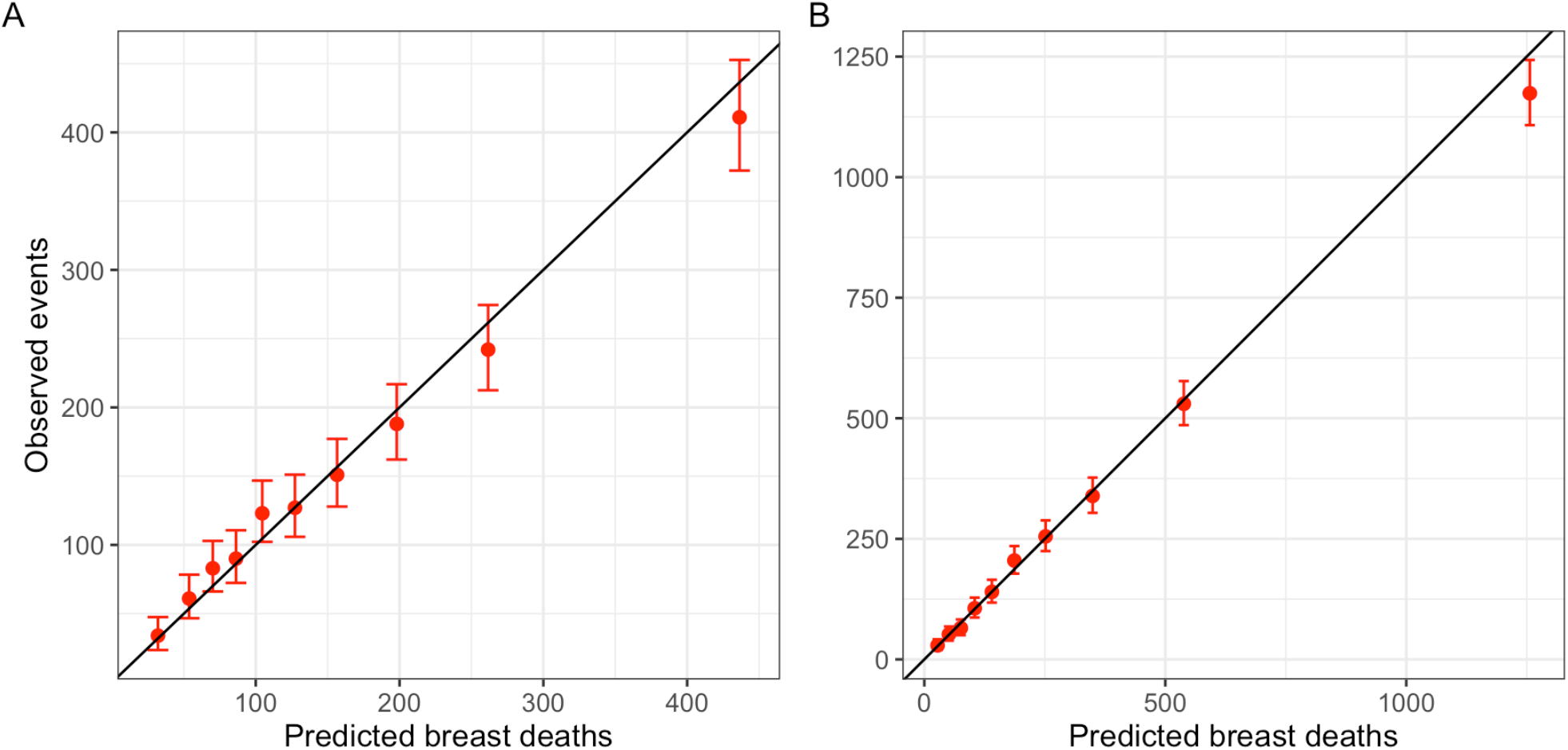
Observed deaths from breast cancer compared to predicted in tenths of predicted risk. A. ER negative cases. B. ER positive cases.

## Discussion

The PREDICT breast prognostic model has been refined and improved over a continuous programme of model development followed by deployment of the model to the PREDICT breast web tool. The major limitations of this study were the incompleteness of data on tumour markers HER2, KI67 and PR and the incomplete data on type of systemic treatment and duration of hormone therapy received by patients. However, the same limitations affected the development of previous versions of the model.

Further development and refinement of the model might include the incorporation of tumour genomic risk scores and prognostic scores based on novel artificial intelligence algorithms applied to routing heamatoxylin and eosin stained pathology sections.

PREDICT breast v4.0 provides a small improvement compared to v3.1 in a contemporary cohort of female breast cancer cases representative of the population in England. Given the good performance of earlier versions of PREDICT breast in other populations we expect v4.0 to perform equally well.

## Data availability

The data used for these analyses cannot be shared by the authors for reasons of confidentiality. They are available on request from the England National Disease Registration Service at https://digital.nhs.uk/services/national-disease-registration-service#requests-for-access-to-ndrs-data.

## Code Availability

All analyses were carried out using the *mfp* ^24^, *patchwork* ^25^, *pROC*,^26^ *survival* ^27^, *tableone* ^28^ and *tidyverse* ^29^ packages for the R software ^30^ implemented in R Studio ^31^. The R markdown script used to analyse the data and draw the figures are available at https://github.com/paul-pharoah/predict.

## Author contributions

PDPP conceived the project carried out the data analysis and wrote the first draft of the manuscript

Y-WH provided feedback on the analyses and edited the manuscript

GCW conceived the project and edited the manuscript.

P-CP provided feedback on the analyses and edited the manuscript

All authors approved the final version of the manuscript.

